# Maternal immune activation induces methylation changes in schizophrenia genes

**DOI:** 10.1101/2022.01.11.22268935

**Authors:** Thomas Johnson, Defne Saatci, Lahiru Handunnetthi

**Author notes:** Correspondence to: Dr Lahiru Handunnetthi, Clinical Lecturer in Neurology, Wellcome Centre for Human Genetics, University of Oxford, OX3 9DU, United Kingdom.

## Abstract

Susceptibility to schizophrenia is mediated by genetic and environmental risk factors. Infection driven maternal immune activation (MIA) during pregnancy is a key environmental risk factor. However, little is known about how MIA during pregnancy could contribute to adult-onset schizophrenia. In this study, we investigated if maternal immune activation induces changes in methylation of genes linked to schizophrenia. We found that differentially expressed genes in schizophrenia brain were significantly enriched among MIA induced differentially methylated genes in the foetal brain in a cell-type-specific manner. Upregulated genes in layer V pyramidal neurons were enriched among hypomethylated genes at gestational day 9 (fold change = 1.57, FDR = 0.049) and gestational day 17 (fold change = 1.97, FDR = 0.0006). We also found that downregulated genes in GABAergic Rosehip interneurons were enriched among hypermethylated genes at gestational day 17 (fold change = 1.62, FDR= 0.03). Collectively, our results highlight a connection between MIA driven methylation changes during gestation and schizophrenia gene expression signatures in the adult brain. These findings carry important implications for early preventative strategies in schizophrenia.

## INTRODUCTION

Epidemiological studies show overwhelming evidence that both genetic and environmental risk factors are important in the aetiology of schizophrenia (Owen et al., 2016). Recently, genome-wide association studies (GWAS) and transcriptomic studies have driven an unprecedented leap in our understanding of the disease, identifying hundreds of genetic variants associated with schizophrenia, as well as gene expression signatures in the adult schizophrenia brain (Consortium et al., 2020; Gandal et al., 2018; Ruzicka et al., 2020). The expression signatures in the schizophrenia brains are cell-type-specific and pathway analyses of these genes highlight the importance of neurodevelopmental processes in the aetiology of schizophrenia (Ruzicka et al., 2020).

Although environmental risk factors are multifaceted, epidemiological associations between maternal exposure to infections in gestation and subsequent increased risk of schizophrenia are established (Brown et al., 2005, 2004; Buka et al., 2008; Mortensen et al., 2007; Saatci et al., 2021; Sorensen et al., 2009). This association has been studied using maternal immune activation (MIA) models in which exposure of pregnant mice to immune insults give rise to disease relevant pathological and behavioural changes in their adult offspring (Gumusoglu and Stevens, 2019). MIA can be achieved by many immune agents such as polyinosinic:polycytidylic acid (polyI:C) and lipopolysaccharides (LPS) mimicking viral and bacterial infections respectively. A recent study showed that MIA induction through polyI:C results in the deregulation of known schizophrenia genes in the foetal brain (Handunnetthi et al., 2021). Despite extensive modelling in animals, our understanding of the how MIA could contribute to long lasting changes in the brain, that increase risk of schizophrenia in the offspring in adulthood, remains poorly understood.

DNA methylation regulates gene expression by recruiting proteins involved in gene repression or by inhibiting the binding of transcription factor(s) to DNA (Boyes and Bird, 1991; Nan et al., 1998). During development, the pattern of DNA methylation in the genome changes because of a dynamic process involving both de novo DNA methylation and demethylation. Previous studies suggest that MIA during pregnancy can influence this dynamic process, providing a possible mechanism by which infection could influence gene expression in the foetal brain (Basil et al., 2014; Richetto et al., 2017; Tang et al., 2013). Furthermore, DNA methylation changes can persist into adulthood, resulting in long lasting modification of gene expression that may continue to influence neural function in later life (Stevenson et al., 2020). This represents an attractive molecular mechanism that could link environmental exposure in early life such as maternal immune activation to adult-onset schizophrenia.

Therefore, we sought to examine the relationship between MIA-induced changes to the foetal methylome and genes known to play a role in schizophrenia pathogenesis. Specifically, this study investigated whether differentially expressed genes (DEGs) in schizophrenia brains and genes identified through GWAS were significantly enriched among differentially methylated genes (DMGs) in the foetal brains following MIA.

## MATERIALS AND METHODOLOGY

### Differentially methylated genes from MIA mouse model of schizophrenia

Two sets of DMGs in foetal brains were extracted after pregnant mice were exposed to polyI:C either in middle (POL-GD9) gestation or late (POL-GD17) gestation from a previous study (Richetto et al., 2017). In this study, methylation differences were identified through genome-wide capture sequencing at single-base resolution for both POL-GD9 and POL-GD17. There were 2364 and 3361 DMGs (defined as methylation change > 5%, FDR < 0.05) at POL-GD9 and POL-GD17 timepoints respectively. Genes that exhibited both hyper- and hypomethylation changes across different domains of the same gene were excluded (POL-GD9: 47 genes and POL-GD17: 80 genes). Finally, DMGs were converted to human orthologues for downstream analyses using the BioMart database (Durinck et al., 2009).

### Schizophrenia-associated genes from GWAS

Two sets of schizophrenia-associated genes were extracted from the largest GWAS to date comprising of 69,369 patients and 236,642 controls (Consortium et al., 2020). The first set included likely causal genes (*n* = 130) that were identified through multiple fine-mapping approaches. The second set covered a broad group of credible genes (*n* = 643) with some genomic support for their role in the disease.

### Differentially expressed genes in schizophrenia brains

Cell-type-specific gene expression patterns linked to schizophrenia were obtained from a previous single-cell RNA-sequencing study of 24 schizophrenia and 24 age- and sex-matched healthy brains (Ruzicka et al., 2020). The tissue samples were derived from the prefrontal cortex of post-mortem brains with single cell libraries generated using the 10x Genomics Chromium Platform and sequenced using an Illumina NextSeq500 machine. Cell-type-specific gene expression patterns were available for 18 cell types, including excitatory cortical neurons, GABAergic interneurons, astrocytes, oligodendrocyte progenitor cells, microglia and endothelial cells in schizophrenia brains. Overall, there were 1637 upregulated and 2492 downregulated genes in schizophrenia brains compared to controls across all cell types.

### Statistical analysis

We tested if schizophrenia genes (GWAS associated genes and DEGs in schizophrenia brains) were enriched among DMGs using a hypergeometric test. The minimum overlap was set to five and the P-values were adjusted for multiple testing by controlling the false discovery rate (FDR). All gene enrichment analyses were conducted using the xEnricher functions in the R package ‘XGR’ (version 1.1.4).

### Data availability and Ethics

Only publicly available, anonymised summary data was used during this study, and we can provide processed data upon reasonable request to the corresponding author. This study was subject to Oxford University’s Research Integrity and Ethics Policy.

## RESULTS

We found significant enrichment of differentially expressed schizophrenia genes among differentially methylated genes in foetal brain after MIA (*figure 1*). Specifically, 1) upregulated genes in layer IV pyramidal neurons were enriched among hypermethylated genes at POL-GD9 (fold change = 2.02, FDR = 0.037), 2) upregulated genes in layer V pyramidal neurons were enriched among hypomethylated genes at POL-GD9 (fold change = 1.57, FDR = 0.049) and at POL-GD17 (fold change = 1.97, FDR = 0.0006), 3) downregulated genes expressed in GABAergic Rosehip interneurons were enriched among hypermethylated genes at POL-GD17 (fold change = 1.62, FDR= 0.03) and finally 4) upregulated genes in oligodendrocytes were enriched among hypomethylated genes at POLD-GD9 (fold change = 2.45, FDR = 0.0045). The enriched genes are summarised in *supplementary table 1*.

**Figure 1.**
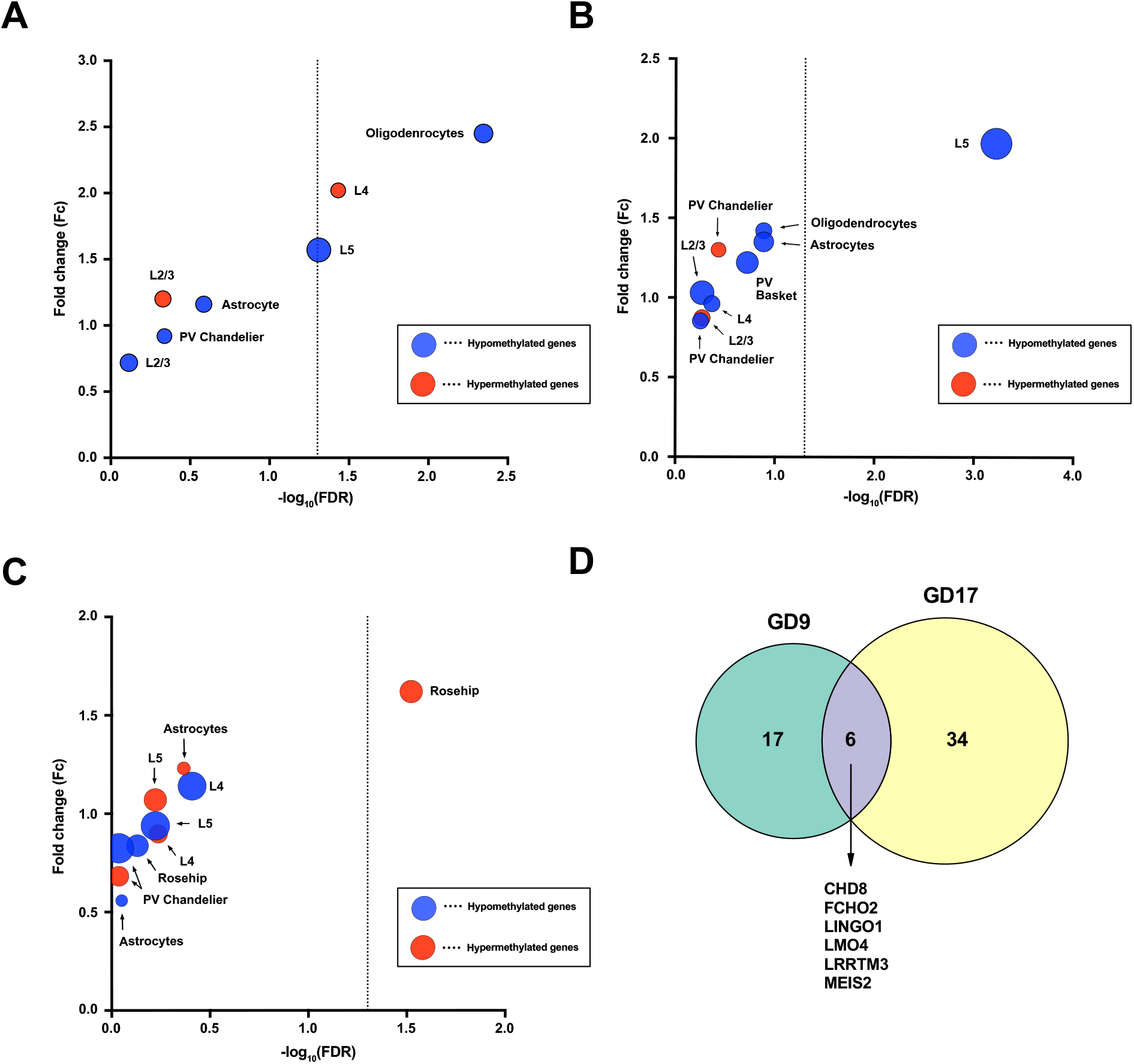
(A) Enrichment of upregulated genes among differentially methylated foetal genes following MIA at GD9. Size of the circles represents the number of overlapping genes. The dashed line is set at FDR = 0.05. (B) Enrichment of upregulated genes among differentially methylated foetal genes following MIA at GD17. Size of the circles represents the number of overlapping genes. The dashed line is set at FDR = 0.05. (C) Enrichment of downregulated genes among differentially methylated foetal genes following MIA at GD17. Sizes of the circles represents the number of overlapping genes. The dashed line is set at FDR = 0.05. (D) The intersect of enriched genes differentially methylated following MIA at both GD9 and GD17.

There was no significant enrichment of GWAS associated genes among differentially methylated genes after MIA at GD9 and GD17, including fine-mapped genes (GD9-hypomethylated: fold change = 0.682, FDR = 0.79; GD17-hypomethylated: fold change = 1.41, FDR = 0.12; GD17-hypermethylated: fold change = 0.806, FDR = 0.59) or genes with broader genomic evidence (GD9-hypomethylated: fold change = 0.41, FDR = 1; GD9-hypermethylated: fold change = 0.423, FDR = 1; GD17-hypomethylated: fold change = 0.653, FDR = 1; GD17-hypermethylated: fold change = 0.479, FDR = 1).

## DISCUSSION

In this study, we demonstrate that MIA can induce methylation changes to known DEGs in schizophrenia brains. The majority of these enriched genes were specific to cortical excitatory and inhibitory neurons, which is in line with the imbalance of excitatory-inhibitory transmission that has long been considered a key feature of schizophrenia (Liu et al., 2021). However, we were unable to detect enrichment of GWAS-associated schizophrenia genes among the DMGs following MIA. This may be because GWAS associated genes represent a narrow group of disease relevant genes that exert their functional effects during neurodevelopment while differentially expressed genes in schizophrenia brains capture many disease processes including those influenced by the environment in schizophrenia pathogenesis. Overall, this work brings together genetic and environmental risk factors in schizophrenia, and sheds light on a potential mechanism that could contribute to the long-term consequences of MIA during gestation.

The enriched genes we identified play key roles in biological processes that are relevant to schizophrenia such as neurodevelopment, synaptic connectivity, and mitochondrial function (summarised in *supplementary table 2*) (OBI-NAGATA et al., 2019; Rajasekaran et al., 2015; Rapoport et al., 2012). For example, we found that *PRKCE* was hypermethylated following MIA. The expression of *PRKCE* is important for neurite outgrowth in response to interaction with neuron growth factor. Consistent with our own results, *PRKCE* was found to be hypermethylated in the dorsolateral prefrontal cortex of schizophrenia brains post-mortem by a DNA methylation assay (Alelú-Paz et al., 2016). Similarly, *HIVEP2*, a transcriptional regulator involved in controlling the activity of many genes implicated in brain development including *SSTR-2, c-Myc*, and genes in the NF-κB pathway (Dörflinger et al., 1999; Fukuda et al., 2002; Iwashita et al., 2012; Wu, 2002), was hypermethylated following MIA. Furthermore, loss-of-function variants in *HIVEP2* are strongly associated with intellectual disability and developmental delay (Srivastava et al., 2016; Steinfeld et al., 2016).

In our study, the methylation data was extracted from mice exposed to PolyI:C during gestation periods corresponding to the end of the first trimester (GD9) and late phase of the second trimester (GD17) in humans (Richetto et al., 2017). We found enrichment of DEGs in schizophrenia brains that were specific to these gestational time points. Evidence from previous studies suggest genes involved in GABAergic neuronal development appear to be influenced to a greater extent by MIA in late gestation (Bitanihirwe et al., 2010; Meyer et al., 2008; Suchiman et al., 2015; Winter et al., 2009). Consistent with this, we only found differentially expressed genes from cortical GABAergic interneurons to be enriched in DMGs following MIA in late gestation. On the contrary, Wnt signalling, which has a crucial role in neurodevelopment, appears to be primarily affected by MIA in early gestation (Hoseth et al., 2018; Richetto et al., 2017). In line with this, we found enrichment of genes known to be involved in Wnt signalling, including *CCND3* and *NXN*, among DMGs following MIA in early gestation.

Our study has several limitations. The *a priori* decision to remove dually hypo- and hypermethylated genes may have resulted in the exclusion of genes pertinent to schizophrenia. Further analysis is required to investigate the effect of MIA and DNA methylation in these genes. Also, our analysis was limited by the quality of the available data, including DMGs from the MIA models, the GWAS-associated genes, and DEGs extracted from the single-cell RNA sequencing study. Further, there are clear translational differences between mice and humans which impedes the ability of polyI:C to fully recapitulate the complexity of immune responses during pregnancy. Finally, we only investigated MIA-induced gene expression changes in one model of MIA mimicking viral infection. Therefore, further work is needed to understand how other models of MIA involving various immune agents could affect the methylation of schizophrenia genes.

In summary, this study provides novel insights into the epigenetic mechanisms underpinning the interplay between genetic and environmental risk factors in the aetiology of schizophrenia. Our results suggest that MIA can induce stable DNA methylation changes that could lead to cell-type-specific gene expression changes in schizophrenia brains. These genes are linked to abnormal neurodevelopment, impaired synaptic connectivity, and mitochondrial dysfunction. Importantly, these findings carry clear implications for disease prevention strategies in schizophrenia because treatment and/or prevention of maternal infection during pregnancy are tractable health goals.

## Data Availability

Only publicly available, anonymised summary data was used during this study, and we can provide processed data upon reasonable request to the corresponding author. This study was subject to Oxford Universitys Research Integrity and Ethics Policy.

https://www.medrxiv.org/content/early/2020/09/13/2020.09.12.20192922.external-links

https://www.medrxiv.org/content/early/2020/11/09/2020.11.06.20225342.external-links

https://www.sciencedirect.com/science/article/pii/S0006322316326713#s0090

## Author statement

TJ: Methodology, Data curation, Formal analysis, Writing – original draft; Writing – review & editing; DS: Writing – original draft; Writing – review & editing; LH: Conceptualization, Funding acquisition, Methodology, Writing – original draft; Writing – review & editing

## REFERENCES

Alelú-Paz, R., Carmona, F.J., Sanchez-Mut, J. V., Cariaga-Martínez, A., González-Corpas, A., Ashour, N., Orea, M.J., Escanilla, A., Monje, A., Guerrero Márquez, C., Saiz-Ruiz, J., Esteller, M., Ropero, S., 2016. Epigenetics in Schizophrenia: A Pilot Study of Global DNA Methylation in Different Brain Regions Associated with Higher Cognitive Functions. Front. Psychol. 7. https://doi.org/10.3389/fpsyg.2016.01496

Basil, P., Li, Q., Dempster, E.L., Mill, J., Sham, P.-C., Wong, C.C.Y., McAlonan, G.M., 2014. Prenatal maternal immune activation causes epigenetic differences in adolescent mouse brain. Transl. Psychiatry 4, e434–e434. https://doi.org/10.1038/tp.2014.80

Bitanihirwe, B.K., Peleg-Raibstein, D., Mouttet, F., Feldon, J., Meyer, U., 2010. Late Prenatal Immune Activation in Mice Leads to Behavioral and Neurochemical Abnormalities Relevant to the Negative Symptoms of Schizophrenia. Neuropsychopharmacology 35, 2462–2478. https://doi.org/10.1038/npp.2010.129

Boyes, J., Bird, A., 1991. DNA methylation inhibits transcription indirectly via a methyl-CpG binding protein. Cell 64, 1123–1134. https://doi.org/10.1016/0092-8674(91)90267-3

Brown, A.S., Begg, M.D., Gravenstein, S., Schaefer, C.A., Wyatt, R.J., Bresnahan, M., Babulas, V.P., Susser, E.S., 2004. Serologic Evidence of Prenatal Influenza in the Etiology of Schizophrenia. Arch. Gen. Psychiatry 61, 774. https://doi.org/10.1001/archpsyc.61.8.774

Brown, A.S., Schaefer, C.A., Quesenberry, C.P., Liu, L., Babulas, V.P., Susser, E.S., 2005. Maternal Exposure to Toxoplasmosis and Risk of Schizophrenia in Adult Offspring. Am. J. Psychiatry 162, 767–773. https://doi.org/10.1176/appi.ajp.162.4.767

Buka, S.L., Cannon, T.D., Torrey, E.F., Yolken, R.H., 2008. Maternal Exposure to Herpes Simplex Virus and Risk of Psychosis Among Adult Offspring. Biol. Psychiatry 63, 809–815. https://doi.org/10.1016/j.biopsych.2007.09.022

Consortium, T.S.W.G. of the P.G., Ripke, S., Walters, J.T.R., O’Donovan, M.C., 2020. Mapping genomic loci prioritises genes and implicates synaptic biology in schizophrenia. medRxiv 2020.09.12.20192922. https://doi.org/10.1101/2020.09.12.20192922

Dörflinger, U., Pscherer, A., Moser, M., Rümmele, P., Schüle, R., Buettner, R., 1999. Activation of Somatostatin Receptor II Expression by Transcription Factors MIBP1 and SEF-2 in the Murine Brain. Mol. Cell. Biol. 19, 3736–3747. https://doi.org/10.1128/MCB.19.5.3736

Durinck, S., Spellman, P.T., Birney, E., Huber, W., 2009. Mapping identifiers for the integration of genomic datasets with the R/Bioconductor package biomaRt. Nat. Protoc. 4, 1184–1191. https://doi.org/10.1038/nprot.2009.97

Fukuda, S., Yamasaki, Y., Iwaki, T., Kawasaki, H., Akieda, S., Fukuchi, N., Tahira, T., Hayashi, K., 2002. Characterization of the Biological Functions of a Transcription Factor, c-mycIntron Binding Protein 1 (MIBP1). J. Biochem. 131, 349–357. https://doi.org/10.1093/oxfordjournals.jbchem.a003109

Gandal, M.J., Zhang, P., Hadjimichael, E., Walker, R.L., Chen, C., Liu, S., Won, H., van Bakel, H., Varghese, M., Wang, Y., Shieh, A.W., Haney, J., Parhami, S., Belmont, J., Kim, M., Moran Losada, P., Khan, Z., Mleczko, J., Xia, Y., Dai, R., Wang, D., Yang, Y.T., Xu, M., Fish, K., Hof, P.R., Warrell, J., Fitzgerald, D., White, K., Jaffe, A.E., Peters, M.A., Gerstein, M., Liu, C., Iakoucheva, L.M., Pinto, D., Geschwind, D.H., Ashley-Koch, A.E., Crawford, G.E., Garrett, M.E., Song, L., Safi, A., Johnson, G.D., Wray, G.A., Reddy, T.E., Goes, F.S., Zandi, P., Bryois, J., Jaffe, A.E., Price, A.J., Ivanov, N.A., Collado-Torres, L., Hyde, T.M., Burke, E.E., Kleiman, J.E., Tao, R., Shin, J.H., Akbarian, S., Girdhar, K., Jiang, Yan, Kundakovic, M., Brown, L., Kassim, B.S., Park, R.B., Wiseman, J.R., Zharovsky, E., Jacobov, R., Devillers, O., Flatow, E., Hoffman, G.E., Lipska, B.K., Lewis, D.A., Haroutunian, V., Hahn, C.-G., Charney, A.W., Dracheva, S., Kozlenkov, A., Belmont, J., DelValle, D., Francoeur, N., Hadjimichael, E., Pinto, D., van Bakel, H., Roussos, P., Fullard, J.F., Bendl, J., Hauberg, M.E., Mangravite, L.M., Peters, M.A., Chae, Y., Peng, J., Niu, M., Wang, X., Webster, M.J., Beach, T.G., Chen, C., Jiang, Yi, Dai, R., Shieh, A.W., Liu, C., Grennan, K.S., Xia, Y., Vadukapuram, R., Wang, Y., Fitzgerald, D., Cheng, L., Brown, Miguel, Brown, Mimi, Brunetti, T., Goodman, T., Alsayed, M., Gandal, M.J., Geschwind, D.H., Won, H., Polioudakis, D., Wamsley, B., Yin, J., Hadzic, T., De La Torre Ubieta, L., Swarup, V., Sanders, S.J., State, M.W., Werling, D.M., An, J.-Y., Sheppard, B., Willsey, A.J., White, K.P., Ray, M., Giase, G., Kefi, A., Mattei, E., Purcaro, M., Weng, Z., Moore, J., Pratt, H., Huey, J., Borrman, T., Sullivan, P.F., Giusti-Rodriguez, P., Kim, Y., Sullivan, P., Szatkiewicz, J., Rhie, S.K., Armoskus, C., Camarena, A., Farnham, P.J., Spitsyna, V.N., Witt, H., Schreiner, S., Evgrafov, O. V., Knowles, J.A., Gerstein, M., Liu, S., Wang, D., Navarro, F.C.P., Warrell, J., Clarke, D., Emani, P.S., Gu, M., Shi, X., Xu, M., Yang, Y.T., Kitchen, R.R., Gürsoy, G., Zhang, J., Carlyle, B.C., Nairn, A.C., Li, M., Pochareddy, S., Sestan, N., Skarica, M., Li, Z., Sousa, A.M.M., Santpere, G., Choi, J., Zhu, Y., Gao, T., Miller, D.J., Cherskov, A., Yang, M., Amiri, A., Coppola, G., Mariani, J., Scuderi, S., Szekely, A., Vaccarino, F.M., Wu, F., Weissman, S., Roychowdhury, T., Abyzov, A., 2018. Transcriptome-wide isoform-level dysregulation in ASD, schizophrenia, and bipolar disorder. Science (80-.). 362, 13–22. https://doi.org/10.1126/science.aat8127

Gumusoglu, S.B., Stevens, H.E., 2019. Maternal Inflammation and Neurodevelopmental Programming: A Review of Preclinical Outcomes and Implications for Translational Psychiatry. Biol. Psychiatry 85, 107–121. https://doi.org/10.1016/j.biopsych.2018.08.008

Handunnetthi, L., Saatci, D., Hamley, J.C., Knight, J.C., 2021. Maternal immune activation downregulates schizophrenia genes in the foetal mouse brain. Brain Commun. 3. https://doi.org/10.1093/braincomms/fcab275

Hoseth, E.Z., Krull, F., Dieset, I., Mørch, R.H., Hope, S., Gardsjord, E.S., Steen, N.E., Melle, I., Brattbakk, H.-R., Steen, V.M., Aukrust, P., Djurovic, S., Andreassen, O.A., Ueland, T., 2018. Exploring the Wnt signaling pathway in schizophrenia and bipolar disorder. Transl. Psychiatry 8, 55. https://doi.org/10.1038/s41398-018-0102-1

Iwashita, Y., Fukuchi, N., Waki, M., Hayashi, K., Tahira, T., 2012. Genome-wide Repression of NF-κB Target Genes by Transcription Factor MIBP1 and Its Modulation by O-Linked β-N-Acetylglucosamine (O-GlcNAc) Transferase. J. Biol. Chem. 287, 9887–9900. https://doi.org/10.1074/jbc.M111.298521

Liu, Y., Ouyang, P., Zheng, Y., Mi, L., Zhao, J., Ning, Y., Guo, W., 2021. A Selective Review of the Excitatory-Inhibitory Imbalance in Schizophrenia: Underlying Biology, Genetics, Microcircuits, and Symptoms. Front. Cell Dev. Biol. 9. https://doi.org/10.3389/fcell.2021.664535

Meyer, U., Nyffeler, M., Yee, B.K., Knuesel, I., Feldon, J., 2008. Adult brain and behavioral pathological markers of prenatal immune challenge during early/middle and late fetal development in mice. Brain. Behav. Immun. 22, 469–486. https://doi.org/10.1016/j.bbi.2007.09.012

Mortensen, P.B., Norgaard-Pedersen, B., Waltoft, B.L., Sorensen, T.L., Hougaard, D., Yolken, R.H., 2007. Early Infections of Toxoplasma gondii and the Later Development of Schizophrenia. Schizophr. Bull. 33, 741–744. https://doi.org/10.1093/schbul/sbm009

Nan, X., Ng, H.-H., Johnson, C.A., Laherty, C.D., Turner, B.M., Eisenman, R.N., Bird, A., 1998. Transcriptional repression by the methyl-CpG-binding protein MeCP2 involves a histone deacetylase complex. Nature 393, 386–389. https://doi.org/10.1038/30764

Obi-Nagata, K., Temma, Y., Hayashi-Takagi, A., 2019. Synaptic functions and their disruption in schizophrenia: From clinical evidence to synaptic optogenetics in an animal model. Proc. Japan Acad. Ser. B 95, 179–197. https://doi.org/10.2183/pjab.95.014

Owen, M.J., Sawa, A., Mortensen, P.B., 2016. Schizophrenia. Lancet 388, 86–97. https://doi.org/10.1016/S0140-6736(15)01121-6

Rajasekaran, A., Venkatasubramanian, G., Berk, M., Debnath, M., 2015. Mitochondrial dysfunction in schizophrenia: Pathways, mechanisms and implications. Neurosci. Biobehav. Rev. 48, 10–21. https://doi.org/10.1016/j.neubiorev.2014.11.005

Rapoport, J.L., Giedd, J.N., Gogtay, N., 2012. Neurodevelopmental model of schizophrenia: update 2012. Mol. Psychiatry 17, 1228–1238. https://doi.org/10.1038/mp.2012.23

Richetto, J., Massart, R., Weber-Stadlbauer, U., Szyf, M., Riva, M.A., Meyer, U., 2017. Genome-wide DNA Methylation Changes in a Mouse Model of Infection-Mediated Neurodevelopmental Disorders. Biol. Psychiatry 81, 265–276. https://doi.org/10.1016/j.biopsych.2016.08.010

Ruzicka, W.B., Mohammadi, S., Davila-Velderrain, J., Subburaju, S., Tso, D.R., Hourihan, M., Kellis, M., 2020. Single-cell dissection of schizophrenia reveals neurodevelopmental-synaptic axis and transcriptional resilience. medRxiv 2020.11.06.20225342. https://doi.org/10.1101/2020.11.06.20225342

Saatci, D., van Nieuwenhuizen, A., Handunnetthi, L., 2021. Maternal infection in gestation increases the risk of non-affective psychosis in offspring: a meta-analysis. J. Psychiatr. Res. 139, 125–131. https://doi.org/10.1016/j.jpsychires.2021.05.039

Sorensen, H.J., Mortensen, E.L., Reinisch, J.M., Mednick, S.A., 2009. Association Between Prenatal Exposure to Bacterial Infection and Risk of Schizophrenia. Schizophr. Bull. 35, 631–637. https://doi.org/10.1093/schbul/sbn121

Srivastava, S., Engels, H., Schanze, I., Cremer, K., Wieland, T., Menzel, M., Schubach, M., Biskup, S., Kreiß, M., Endele, S., Strom, T.M., Wieczorek, D., Zenker, M., Gupta, S., Cohen, J., Zink, A.M., Naidu, S., 2016. Loss-of-function variants in HIVEP2 are a cause of intellectual disability. Eur. J. Hum. Genet. 24, 556–561. https://doi.org/10.1038/ejhg.2015.151

Steinfeld, H., Cho, M.T., Retterer, K., Person, R., Schaefer, G.B., Danylchuk, N., Malik, S., Wechsler, S.B., Wheeler, P.G., van Gassen, K.L.I., Terhal, P.A., Verhoeven, V.J.M., van Slegtenhorst, M.A., Monaghan, K.G., Henderson, L.B., Chung, W.K., 2016. Mutations in HIVEP2 are associated with developmental delay, intellectual disability, and dysmorphic features. Neurogenetics 17, 159–164. https://doi.org/10.1007/s10048-016-0479-z

Stevenson, K., Lillycrop, K.A., Silver, M.J., 2020. Fetal programming and epigenetics. Curr. Opin. Endocr. Metab. Res. 13, 1–6. https://doi.org/10.1016/j.coemr.2020.07.005

Suchiman, H.E.D., Slieker, R.C., Kremer, D., Slagboom, P.E., Heijmans, B.T., Tobi, E.W., 2015. Design, measurement and processing of region-specific DNA methylation assays: the mass spectrometry-based method EpiTYPER. Front. Genet. 6. https://doi.org/10.3389/fgene.2015.00287

Tang, B., Jia, H., Kast, R.J., Thomas, E.A., 2013. Epigenetic changes at gene promoters in response to immune activation in utero. Brain. Behav. Immun. 30, 168–175. https://doi.org/10.1016/j.bbi.2013.01.086

Winter, C., Djodari-Irani, A., Sohr, R., Morgenstern, R., Feldon, J., Juckel, G., Meyer, U., 2009. Prenatal immune activation leads to multiple changes in basal neurotransmitter levels in the adult brain: implications for brain disorders of neurodevelopmental origin such as schizophrenia. Int. J. Neuropsychopharmacol. 12, 513. https://doi.org/10.1017/S1461145708009206

Wu, L.-C., 2002. ZAS: C2H2 Zinc Finger Proteins Involved in Growth and Development. Gene Expr. 10, 137–152. https://doi.org/10.3727/000000002783992479

