## Supplemental Table 1 for "Maternal immune activation induces methylation changes in schizophrenia genes"

*Supplementary Table 1*

| **Cell Type** | **Gestation Day** | **Methylation Status** | **DEGs** | **Number overlapping** | **Overlapping Gene Names** |
| --- | --- | --- | --- | --- | --- |
| L4 | 9 | Hypermethylated | Upregulated | 5 | *ADAMTS19, CCND3, HSPB1, PDZRN4, TLE4* |
| L5 | 9 | Hypomethylated | Upregulated | 13 | *CDH8, CNTN1, FCHO2, LINGO1, LMO4, LRRTM3, MEIS2, NXN, RARB, ROBO1, TCF4, TENM2, TPD52L1* |
| L5 | 17 | Hypomethylated | Upregulated | 22 | *ANO6, APP, BNIP3L, CDH8, EFR3A, FCHO2, IQCJ-SCHIP1, LINGO1, LMO4, LRRTM3, MAPK10, MAPRE2, MBNL1, MEIS2, NEK7, OSBPL1A, PELI2, PTPRU, RAPGEF2, RPRM, SNX31, TJP1* |
| Rosehip | 17 | Hypermethylated | Downregulated | 18 | *ANKRD10, DHDDS, DOK5, EVA1C, FBXW4, FNIP2, FRAS1, GABPB2, GAS8, HIVEP1, MICU1, PDE10A, PHC1, PRKCE, RAB11FIP4, ST3GAL1, SV2C, TANGO6* |
| Oli | 9 | Hypomethylated | Upregulated | 8 | *L3MBTL4, MEIS2, NFASC, NFIB, ROBO1, SLC38A2, TANC1, TCF4* |
