## Supplemental Table 2 for "Maternal immune activation induces methylation changes in schizophrenia genes"

*Supplementary Table* *2*

| **Gene Name** | **Cell Type Enrichment** | **Description** |
| --- | --- | --- |
| *Impaired Neurodevelopment* | | |
| RAPGEF2 | Cortical layer V pyramidal neurons | RAPGEF2 encodes guanine nucleotide exchange factors necessary for the expression of apical adheren junction proteins, which are important in neurodevelopment and have been implicated in schizophrenia pathogenesis (O’Dushlaine et al., 2011; Veeraval et al., 2020). Abnormal RAPGEF2 expression results in dysregulated neurogenesis and aberrant radial glial cell migration (Farag et al., 2017; Maeta et al., 2016). Furthermore, several sequencing analyses have shown rare copy number variants encompassing RAPGEF2 to be significantly associated with sporadic and familial schizophrenia (Xu et al., 2009, 2008). |
| CNTN1 | Cortical layer V pyramidal neurons | CNTN1 encodes an adhesive protein, known as contactin-1/F3 (Cntn1) belonging to the immunoglobulin family and is predominantly expressed in the nervous system (Shimoda and Watanabe, 2009). Interactions between Cntn1 and its ligands are important for neural cell adhesion, myelination, neurite growth, axonal elongation, and fasciculation (Falk et al., 2002; Karagogeos, 2003). *Cntn1* knockdown in neural stem cells resulted in a delay in neuronal migration and abnormal morphology (Chen et al., 2018). Furthermore, mutations in the related CNTNAP2 gene have been associated with schizophrenia and autism spectrum disorder (Bakkaloglu et al., 2008; Friedman et al., 2008). |
| LRRTM3 | Cortical layer V pyramidal neurons | Leucine-rich repeat (LRR)-containing transmembrane proteins (LRRTM) are a family of synaptogenic adhesion molecules implicated in synaptogenesis (Ko, 2012). Overexpression of LRRTM3 has been shown to increase excitatory synapse density in dentate gyrus granule neurons. Further, LRRTM3 control appears to be necessary for activity-regulated AMPA receptor surface expression (Um et al., 2016). Deficiencies in other LRRTM proteins, such as *Lrrtm1*, result in cognitive deficiencies and abnormal hippocampal morphology in mice (Takashima et al., 2011). There is also evidence for the role of LRRTM1 in the pathogenesis of schizophrenia (Francks et al., 2007; Ludwig et al., 2009). |
| ST3GAL1 | Rosehip interneurons | A meta-analysis of 2 GWAS schizophrenia studies identified ST3GAL1 as being significantly associated with negative symptoms in schizophrenia (Xu et al., 2013). This is compelling because another ST3-related gene, ST8SIA2, codes for another nerve cell adhesion molecule (Ono et al., 1994), and appears to play a key role in cell-cell interaction in the developing brain and has previously been described as having an association with schizophrenia risk (Arai et al., 2006). Gene Ontology analysis showed that GO categories associated with ST3GAL1, including O-Glycan biosynthesis and glycosphingolipid biosynthesis, paralleled that of ST8SIA2 (Xu et al., 2013). Furthermore, previous studies have found abnormal levels of N-glycan products in the CSF and serum of patients with schizophrenia, as well as decreased expression of enzymes involved in glycan synthesis in schizophrenia brain tissue (Narayan et al., 2009; Stanta et al., 2010). |
| FRAS1 | Rosehip interneurons | FRAS1 encodes a basement membrane protein diffusely localised throughout the foetal mouse brain. *Fras1* deficient mice exhibit abnormal behavioural phenotypes, including impaired learning and working memory, as well as severely disorganised perineuronal networks in cortical areas (Kalpachidou et al., 2021). In humans, FRASER1 mutations result in a rare development disorder characterised by severe physical malformations and mental retardation, supporting the essential role of FRASER1 expression in development (Dumitru et al., 2016). |
| LINGO-1 | Cortical layer V pyramidal neurons | LINGO-1 is a potential negative regulator of axonal myelination and neurite extension during neurogenesis (Mi et al., 2005; Zhang et al., 2009). In support of our findings, a post-mortem examination of the dorsolateral prefrontal cortex in schizophrenic patients showed increased Lingo-1 expression (Fernandez-Enright et al., 2014). Furthermore, missense variants of LINGO-1 are a reported cause of intellectual disability in humans (Ansar et al., 2018). |
| IQCJ-SCHIP1 | Cortical layer V pyramidal neurons | IQCJ-SCHIP1 encodes a cytoplasmic cytoskeleton-associated protein that binds to cell adhesion molecules and has an important role in axogenesis during development. *Schip1* mutant mice exhibit deficits in anterior commissure development due to impaired axon guidance (Klingler et al., 2015). Impaired interhemispheric connectivity via the anterior commissure has also been observed in schizophrenia patients (Choi et al., 2011). Furthermore, a recessive point mutation in IQCJ-SHIP1 has also been linked to a rare brain malformation syndrome characterised by macroscopic anatomical abnormalities, such as thinning of the corpus callosum, and impaired cognition (Elsaid et al., 2018). |
| TENM2 | Cortical layer V pyramidal neurons | Whole genome sequencing of 61 individuals from multiple schizophrenia families identified TENM2 variants as having significant disease association (Li et al., 2021). TENM2 encodes for Teneurin-2, which plays a role in synaptogenesis, neurite outgrowth, axon guidance, and neuronal connectivity (Silva et al., 2011). |
| PRKCE | Rosehip interneurons | PRKCE gene expression has been found to promote neurite outgrowth in response to their interaction with neuron growth factor. Consistent with our own results, PRKCE was found to be hypermethylated in the dorsolateral prefrontal cortex of schizophrenia brains post-mortem by a DNA methylation assay (Alelú-Paz et al., 2016). In humans, an association study of schizophrenia candidate genes and genes with GO terms relating to neurodevelopment highlighted a cluster of SNPs residing in PRKCE (Zhao et al., 2013). Hence, repressed PRKCE expression may also contribute to dysregulated neurogenesis during foetal development. |
| NFASC | Oligodendrocytes | NFASC encodes for neurofascin, an immunoglobulin cell adhesion molecule important in axogenesis and synapse formation during nervous system development (Ghosh et al., 2018). NFASC deficits have been shown to reduce size and amount of neurite outgrowth in developing neurons (Kvarnung et al., 2019). Furthermore, abnormal NFASC expression has been reported in the superior temporal gyrus of schizophrenia brains (Roussos, 2012). NFASC variants have also been associated with a rare neurodevelopmental disorder characterised by both peripheral and central neurological symptoms (Efthymiou et al., 2019). |
| CHD8 | Cortical layer V neurons | CHD8, encoding an ATP-dependent chromatin-remodelling factor, was found to be hypermethylated at GD17 and enriched amongst upregulated genes expressed by cortical layer V neurons. Suppressed CHD8 expression in healthy induced pluripotent stem cell (iPSC)-derived neural progenitor cells resulted in the modulation of various gene pathways related to neurodevelopment (Sugathan et al., 2014). This was also revalidated by CRISPR-Cas9-mediated knockout of CHD8 in an iPSC-derived organoid (Wang et al., 2017). Several studies have identified CDH8 variants as significant risk factors for autism spectrum disorder (ASD), a neurodevelopmental disorder whose aetiology is believed to share a common molecular origin with schizophrenia (Krumm et al., 2014; McCarthy et al., 2014; Talkowski et al., 2012). Indeed, a large exome-sequencing study identified CHD8-R2333C mutation in a single case of schizophrenia in a 57yr old female, who also displayed features of ASD (Kimura et al., 2016). |
| TCF4 | Cortical layer V pyramidal neurons  Oligodendrocytes | TCF4 is known to be important for the development and terminal differentiation of oligodendrocytes (Phan et al., 2020; Wedel et al., 2020). TCF4 polymorphisms have also been associated with many psychiatric disorders, including schizophrenia (Teixeira et al., 2021). Furthermore, TCF4 mutations cause a devastating autism spectrum disorder known as Pitts-Hopkins Syndrome, characterised by severe intellectual disability and delayed development (Li et al., 2019; Quednow et al., 2014). An analysis of the genomic locations of TCF4 binding sites showed significant overlap with known genetic risk factors for schizophrenia (Forrest et al., 2018). Furthermore, TCF4 is expressed at increased levels in the prefrontal cortex of schizophrenia brains, consistent with our results suggesting that hypomethylation, and subsequent gene activation, may lead to long-term upregulation of TCF4 into adulthood (Ma et al., 2018). |
| LMO4 | Cortical layer V pyramidal neurons | LMO4 encodes a transcriptional regulator necessary for normal patterns of proliferation and for survival of neural epithelial cells in the neural tube (Lee et al., 2005). Glutamatergic neuron-selective ablation of *Lmo4* leads to schizophrenia-like behaviours in mice, such as impaired working memory and defective sensory gating (Qin et al., 2020). *Lmo4* ablation has also been shown to reduce endocannabinoid production from metabotropic glutamate receptors, which is intriguing given the known association between cannabis usage and schizophrenia-symptoms (Qin et al., 2015). |
| HIVEP2 | Rosehip interneurons | HIVEP2 encodes a large transcriptional regulator involved in regulating the activity of various genes, many of which are implicated in brain development, including SSTR-2, c-Myc, and genes in the NF-κB pathway (Dörflinger et al., 1999; Fukuda et al., 2002; Iwashita et al., 2012; Wu, 2002). Furthermore, loss-of-function variants in HIVEP2 are strongly associated with intellectual disability and developmental delay (Srivastava et al., 2016; Steinfeld et al., 2016). |
| PHC1 | Rosehip interneurons | PHC1 encodes a subunit of the PcG PRC1 complex, which is involved in chromatin remodelling and histone modification, and implicated is in various stages of neurodevelopment (Corley and Kroll, 2015). Furthermore, a bioinformatic analysis of sequencing data extracted from a 22q11.2 deletion syndrome family identified PHC1 variants as significant ‘second hits’ associated with the psychotic phenotype, likely because of their interaction with the 22q11.2 haploinsufficiency (Michaelovsky et al., 2019). |
| CCND3 | Cortical layer IV pyramidal neurons | CCND3 is a key driver of the cell cycle and involved in downstream signalling of the Wnt pathway, which is known to be important in neurodevelopment (Meffre et al., 2014; Valvezan and Klein, 2012). Canonical Wnt signalling has been found to be attenuated in schizophrenia brains (Hoseth et al., 2018). It is possible that the upregulated CCND3 gene expression that we observed in cortical layer IV reflects the outcome of other related MIA-induced epigenetic changes attenuating the Wnt signalling pathway. Consistent with this idea, previous studies have demonstrated that CCND3 expression is increased in response to impaired Wnt signalling (Keats et al., 2014; Tanaka et al., 2011). |
| NXN | Cortical layer V pyramidal neurons | NXN encodes a redox-dependent negative regulator of the Wnt signalling (Funato et al., 2006). Wnt signalling appears to be dysregulated in schizophrenia and may contribute to abnormal neurodevelopment (Hoseth et al., 2018). |
| RARB | Cortical layer V pyramidal neurons | Retinoid metabolites of Vitamin A are important in neurodevelopment and have been linked to the pathogenesis of schizophrenia (Goodman, 1998; Lerner et al., 2016). A large mega-analysis of schizophrenia risk variants involving a GWAS identified five genes involved in retinoid biology are located in genome-wide significant loci (“Biological insights from 108 schizophrenia-associated genetic loci,” 2014). Furthermore, cases of schizophrenia with severe cognitive deficits were found to be enriched in rare variation in the retinoic acid receptor beta gene RARB (Reay et al., 2020), suggesting that disrupted retinoid signalling may be involved in schizophrenia. |
| *Impaired Synaptic Function* | | |
| EFR3A | Cortical layer V pyramidal neurons | EFR3A encodes a protein complex essential for phosphoinositide synthesis at the neural synapse; phosphoinositide’s are integral to subcellular processes at the neural synapse, including membrane transport, cytoskeletal function and plasma membrane signalling (Raghu et al., 2019). Rare non-synonymous mutations in ERF3A were found to be significantly more common in patients with ASD and co-localised with the expression of ASD-related genes (Gupta et al., 2014). |
| SNX31 | Cortical layer V pyramidal neurons | SNX31 expression regulates endocytic vesicle trafficking at neuronal synapses necessary for synaptic transmission (Vieira et al., 2021). A whole exome sequencing study comparing 2 individuals with 22q11.2 deletion, one with and one without psychosis, identified a frameshift mutation in SNX31 as a potential causal variant of the psychotic phenotype (Balan et al., 2014). The potential role of SNX31 in schizophrenia pathogenesis is compelling as other members of the SNX protein family, such as SNX19, are known to be influenced by known genetic risk variants via epigenetic mechanisms and differentially expressed in schizophrenia brains (Ma et al., 2020). |
| *Impaired Mitochondrial Function* | | |
| MICU1 | Rosehip interneurons | MICU1 encodes a calcium transporter that relays cytoplasmic fluctuations in Ca2+ to the mitochondrial matrix to control vital functions like ATP production but can also overload mitochondria with Ca^2+^ to promote cell death (De Stefani et al., 2011; Perocchi et al., 2010). Recently, a rare genetic disorder characterised by learning disabilities and proximal myopathy has been attributed to a MICU1 mutation (Logan et al., 2014), suggesting that altering MICU1 expression could also influence neurodevelopment during the foetal period. |
| BNIP3L | Cortical layer V pyramidal neurons | BNIP3L and encodes a mitochondrial protein involved in promoting and sustaining basal mitophagy (Gao et al., 2015; Shi et al., 2014). In 2017, a GWAS identified 30 new schizophrenia susceptibility loci around BNIP3L (Li et al., 2017). Subsequently, a sequencing of exons and untranslated regions of the BNIP3L locus in patients with schizophrenia revealed three rare non-synonymous mutations occurring at a significantly higher frequency than in healthy controls (Zhou et al., 2020). Our results support the potential role of aberrant BNIP3L expression and abnormal mitophagy in the pathogenesis of schizophrenia, although further functional verification is required. |
| *Neuroinflammation* | | |
| HSPB1 | Cortical layer IV pyramidal neurons | Neuroinflammation is a well-established biomarker in a subset of schizophrenia patients and is driven by activation of the NFkB signalling pathway (Murphy et al., 2021). The NFkB signalling pathway is, in turn, regulated by the expression of HSPB1, which we found to be upregulated in cortical layer IV neurons. Transgenic mice overexpressing human HSPB1 protein show enhanced neuroinflammatory markers following ethanol treatment (Dukay et al., 2021). Alcohol misuse is an established risk factor for psychosis (Nielsen et al., 2017), and it’s possible that susceptible individuals overexpress HSPB1 in adulthood, thereby driving an exaggerated, chronic neuroinflammatory response to environmental insults such as alcohol exposure. |

**SUPPLEMENTARY REFERENCES**

Alelú-Paz, R., Carmona, F.J., Sanchez-Mut, J. V., Cariaga-Martínez, A., González-Corpas, A., Ashour, N., Orea, M.J., Escanilla, A., Monje, A., Guerrero Márquez, C., Saiz-Ruiz, J., Esteller, M., Ropero, S., 2016. Epigenetics in Schizophrenia: A Pilot Study of Global DNA Methylation in Different Brain Regions Associated with Higher Cognitive Functions. Front. Psychol. 7. https://doi.org/10.3389/fpsyg.2016.01496

Ansar, M., Riazuddin, Saima, Sarwar, M.T., Makrythanasis, P., Paracha, S.A., Iqbal, Z., Khan, J., Assir, M.Z., Hussain, M., Razzaq, A., Polla, D.L., Taj, A.S., Holmgren, A., Batool, N., Misceo, D., Iwaszkiewicz, J., de Brouwer, A.P.M., Guipponi, M., Hanquinet, S., Zoete, V., Santoni, F.A., Frengen, E., Ahmed, J., Riazuddin, Sheikh, van Bokhoven, H., Antonarakis, S.E., 2018. Biallelic variants in LINGO1 are associated with autosomal recessive intellectual disability, microcephaly, speech and motor delay. Genet. Med. 20, 778–784. https://doi.org/10.1038/gim.2017.113

Arai, M., Yamada, K., Toyota, T., Obata, N., Haga, S., Yoshida, Y., Nakamura, K., Minabe, Y., Ujike, H., Sora, I., Ikeda, K., Mori, N., Yoshikawa, T., Itokawa, M., 2006. Association Between Polymorphisms in the Promoter Region of the Sialyltransferase 8B (SIAT8B) Gene and Schizophrenia. Biol. Psychiatry 59, 652–659. https://doi.org/10.1016/j.biopsych.2005.08.016

Bakkaloglu, B., O’Roak, B.J., Louvi, A., Gupta, A.R., Abelson, J.F., Morgan, T.M., Chawarska, K., Klin, A., Ercan-Sencicek, A.G., Stillman, A.A., Tanriover, G., Abrahams, B.S., Duvall, J.A., Robbins, E.M., Geschwind, D.H., Biederer, T., Gunel, M., Lifton, R.P., State, M.W., 2008. Molecular Cytogenetic Analysis and Resequencing of Contactin Associated Protein-Like 2 in Autism Spectrum Disorders. Am. J. Hum. Genet. 82, 165–173. https://doi.org/10.1016/j.ajhg.2007.09.017

Balan, S., Iwayama, Y., Toyota, T., Toyoshima, M., Maekawa, M., Yoshikawa, T., 2014. 22q11.2 deletion carriers and schizophrenia-associated novel variants. Br. J. Psychiatry 204, 398–399. https://doi.org/10.1192/bjp.bp.113.138420

Biological insights from 108 schizophrenia-associated genetic loci, 2014. . Nature 511, 421–427. https://doi.org/10.1038/nature13595

Chen, Y.-A., Lu, I.-L., Tsai, J.-W., 2018. Contactin-1/F3 Regulates Neuronal Migration and Morphogenesis Through Modulating RhoA Activity. Front. Mol. Neurosci. 11. https://doi.org/10.3389/fnmol.2018.00422

Choi, H., Kubicki, M., Whitford, T.J., Alvarado, J.L., Terry, D.P., Niznikiewicz, M., McCarley, R.W., Kwon, J.S., Shenton, M.E., 2011. Diffusion tensor imaging of anterior commissural fibers in patients with schizophrenia. Schizophr. Res. 130, 78–85. https://doi.org/10.1016/j.schres.2011.04.016

Corley, M., Kroll, K.L., 2015. The roles and regulation of Polycomb complexes in neural development. Cell Tissue Res. 359, 65–85. https://doi.org/10.1007/s00441-014-2011-9

De Stefani, D., Raffaello, A., Teardo, E., Szabò, I., Rizzuto, R., 2011. A forty-kilodalton protein of the inner membrane is the mitochondrial calcium uniporter. Nature 476, 336–340. https://doi.org/10.1038/nature10230

Dörflinger, U., Pscherer, A., Moser, M., Rümmele, P., Schüle, R., Buettner, R., 1999. Activation of Somatostatin Receptor II Expression by Transcription Factors MIBP1 and SEF-2 in the Murine Brain. Mol. Cell. Biol. 19, 3736–3747. https://doi.org/10.1128/MCB.19.5.3736

Dukay, B., Walter, F.R., Vigh, J.P., Barabási, B., Hajdu, P., Balassa, T., Migh, E., Kincses, A., Hoyk, Z., Szögi, T., Borbély, E., Csoboz, B., Horváth, P., Fülöp, L., Penke, B., Vígh, L., Deli, M.A., Sántha, M., Tóth, M.E., 2021. Neuroinflammatory processes are augmented in mice overexpressing human heat-shock protein B1 following ethanol-induced brain injury. J. Neuroinflammation 18, 22. https://doi.org/10.1186/s12974-020-02070-2

Dumitru, A., Costache, M., Lazaroiu, A.M., Simion, G., Secara, D., Cirstoiu, M., Emanoil, A., Georgescu, T.A., Sajin, M., 2016. Fraser Syndrome - a Case Report and Review of Literature. Maedica (Buchar). 11, 80–83.

Efthymiou, S., Salpietro, V., Malintan, N., Poncelet, M., Kriouile, Y., Fortuna, S., De Zorzi, R., Payne, K., Henderson, L.B., Cortese, A., Maddirevula, S., Alhashmi, N., Wiethoff, S., Ryten, M., Botia, J.A., Provitera, V., Schuelke, M., Vandrovcova, J., Groppa, S., Karashova, B.M., Nachbauer, W., Boesch, S., Arning, L., Timmann, D., Cormand, B., Pérez-Dueñas, B., Goraya, J.S., Sultan, T., Mine, J., Avdjieva, D., Kathom, H., Tincheva, R., Banu, S., Pineda-Marfa, M., Veggiotti, P., Ferrari, M.D., van den Maagdenberg, A.M.J.M., Verrotti, A., Marseglia, G., Savasta, S., García-Silva, M., Ruiz, A.M., Garavaglia, B., Borgione, E., Portaro, S., Sanchez, B.M., Boles, R., Papacostas, S., Vikelis, M., Rothman, J., Kullmann, D., Papanicolaou, E.Z., Dardiotis, E., Maqbool, S., Ibrahim, S., Kirmani, S., Rana, N.N., Atawneh, O., Lim, S.-Y., Shaikh, F., Koutsis, G., Breza, M., Mangano, S., Scuderi, C., Borgione, E., Morello, G., Stojkovic, T., Zollo, M., Heimer, G., Dauvilliers, Y.A., Minetti, C., Al-Khawaja, I., Al-Mutairi, F., Hamed, S., Pipis, M., Bettencourt, C., Rinaldi, S., Walsh, L., Torti, E., Iodice, V., Najafi, M., Karimiani, E.G., Maroofian, R., Siquier-Pernet, K., Boddaert, N., De Lonlay, P., Cantagrel, V., Aguennouz, M., El Khorassani, M., Schmidts, M., Alkuraya, F.S., Edvardson, S., Nolano, M., Devaux, J., Houlden, H., 2019. Biallelic mutations in neurofascin cause neurodevelopmental impairment and peripheral demyelination. Brain 142, 2948–2964. https://doi.org/10.1093/brain/awz248

Elsaid, M.F., Chalhoub, N., Ben-Omran, T., Kamel, H., AL Mureikhi, M., Ibrahim, K., Elizabeth Ross, M., Abdel Aleem, A.K., 2018. Homozygous nonsense mutation in SCHIP1/IQCJ-SCHIP1 causes a neurodevelopmental brain malformation syndrome. Clin. Genet. 93, 387–391. https://doi.org/10.1111/cge.13122

Falk, J., Bonnon, C., Girault, J.-A., Faivre-Sarrailh, C., 2002. F3/contactin, a neuronal cell adhesion molecule implicated in axogenesis and myelination. Biol. Cell 94, 327–334. https://doi.org/10.1016/S0248-4900(02)00006-0

Farag, M.I., Yoshikawa, Y., Maeta, K., Kataoka, T., 2017. Rapgef2, a guanine nucleotide exchange factor for Rap1 small GTPases, plays a crucial role in adherence junction (AJ) formation in radial glial cells through ERK-mediated upregulation of the AJ-constituent protein expression. Biochem. Biophys. Res. Commun. 493, 139–145. https://doi.org/10.1016/j.bbrc.2017.09.062

Fernandez-Enright, F., Andrews, J.L., Newell, K.A., Pantelis, C., Huang, X.F., 2014. Novel implications of Lingo-1 and its signaling partners in schizophrenia. Transl. Psychiatry 4, e348–e348. https://doi.org/10.1038/tp.2013.121

Forrest, M.P., Hill, M.J., Kavanagh, D.H., Tansey, K.E., Waite, A.J., Blake, D.J., 2018. The Psychiatric Risk Gene Transcription Factor 4 (TCF4) Regulates Neurodevelopmental Pathways Associated With Schizophrenia, Autism, and Intellectual Disability. Schizophr. Bull. 44, 1100–1110. https://doi.org/10.1093/schbul/sbx164

Francks, C., Maegawa, S., Laurén, J., Abrahams, B.S., Velayos-Baeza, A., Medland, S.E., Colella, S., Groszer, M., McAuley, E.Z., Caffrey, T.M., Timmusk, T., Pruunsild, P., Koppel, I., Lind, P.A., Matsumoto-Itaba, N., Nicod, J., Xiong, L., Joober, R., Enard, W., Krinsky, B., Nanba, E., Richardson, A.J., Riley, B.P., Martin, N.G., Strittmatter, S.M., Möller, H.-J., Rujescu, D., St Clair, D., Muglia, P., Roos, J.L., Fisher, S.E., Wade-Martins, R., Rouleau, G.A., Stein, J.F., Karayiorgou, M., Geschwind, D.H., Ragoussis, J., Kendler, K.S., Airaksinen, M.S., Oshimura, M., DeLisi, L.E., Monaco, A.P., 2007. LRRTM1 on chromosome 2p12 is a maternally suppressed gene that is associated paternally with handedness and schizophrenia. Mol. Psychiatry 12, 1129–1139. https://doi.org/10.1038/sj.mp.4002053

Friedman, J.I., Vrijenhoek, T., Markx, S., Janssen, I.M., van der Vliet, W.A., Faas, B.H.W., Knoers, N. V, Cahn, W., Kahn, R.S., Edelmann, L., Davis, K.L., Silverman, J.M., Brunner, H.G., van Kessel, A.G., Wijmenga, C., Ophoff, R.A., Veltman, J.A., 2008. CNTNAP2 gene dosage variation is associated with schizophrenia and epilepsy. Mol. Psychiatry 13, 261–266. https://doi.org/10.1038/sj.mp.4002049

Fukuda, S., Yamasaki, Y., Iwaki, T., Kawasaki, H., Akieda, S., Fukuchi, N., Tahira, T., Hayashi, K., 2002. Characterization of the Biological Functions of a Transcription Factor, c-mycIntron Binding Protein 1 (MIBP1). J. Biochem. 131, 349–357. https://doi.org/10.1093/oxfordjournals.jbchem.a003109

Funato, Y., Michiue, T., Asashima, M., Miki, H., 2006. The thioredoxin-related redox-regulating protein nucleoredoxin inhibits Wnt–β-catenin signalling through Dishevelled. Nat. Cell Biol. 8, 501–508. https://doi.org/10.1038/ncb1405

Gao, F., Chen, D., Si, J., Hu, Q., Qin, Z., Fang, M., Wang, G., 2015. The mitochondrial protein BNIP3L is the substrate of PARK2 and mediates mitophagy in PINK1/PARK2 pathway. Hum. Mol. Genet. 24, 2528–2538. https://doi.org/10.1093/hmg/ddv017

Ghosh, A., Sherman, D.L., Brophy, P.J., 2018. The Axonal Cytoskeleton and the Assembly of Nodes of Ranvier. Neurosci. 24, 104–110. https://doi.org/10.1177/1073858417710897

Goodman, A.B., 1998. Three independent lines of evidence suggest retinoids as causal to schizophrenia. Proc. Natl. Acad. Sci. 95, 7240–7244. https://doi.org/10.1073/pnas.95.13.7240

Gupta, A.R., Pirruccello, M., Cheng, F., Kang, H., Fernandez, T. V, Baskin, J.M., Choi, M., Liu, L., Ercan-Sencicek, A., Murdoch, J.D., Klei, L., Neale, B.M., Franjic, D., Daly, M.J., Lifton, R.P., De Camilli, P., Zhao, H., Šestan, N., State, M.W., 2014. Rare deleterious mutations of the gene EFR3A in autism spectrum disorders. Mol. Autism 5, 31. https://doi.org/10.1186/2040-2392-5-31

Hoseth, E.Z., Krull, F., Dieset, I., Mørch, R.H., Hope, S., Gardsjord, E.S., Steen, N.E., Melle, I., Brattbakk, H.-R., Steen, V.M., Aukrust, P., Djurovic, S., Andreassen, O.A., Ueland, T., 2018. Exploring the Wnt signaling pathway in schizophrenia and bipolar disorder. Transl. Psychiatry 8, 55. https://doi.org/10.1038/s41398-018-0102-1

Iwashita, Y., Fukuchi, N., Waki, M., Hayashi, K., Tahira, T., 2012. Genome-wide Repression of NF-κB Target Genes by Transcription Factor MIBP1 and Its Modulation by O-Linked β-N-Acetylglucosamine (O-GlcNAc) Transferase. J. Biol. Chem. 287, 9887–9900. https://doi.org/10.1074/jbc.M111.298521

Kalpachidou, T., Makrygiannis, A.K., Pavlakis, E., Stylianopoulou, F., Chalepakis, G., Stamatakis, A., 2021. Behavioural effects of extracellular matrix protein Fras1 depletion in the mouse. Eur. J. Neurosci. 53, 3905–3919. https://doi.org/10.1111/ejn.14759

Karagogeos, D., 2003. Neural GPI anchored cell adhesion molecules. Front. Biosci. 8, 1214. https://doi.org/10.2741/1214

Keats, E.C., Dominguez, J.M., Grant, M.B., Khan, Z.A., 2014. Switch from Canonical to Noncanonical Wnt Signaling Mediates High Glucose-Induced Adipogenesis. Stem Cells 32, 1649–1660. https://doi.org/10.1002/stem.1659

Kimura, H., Wang, C., Ishizuka, K., Xing, J., Takasaki, Y., Kushima, I., Aleksic, B., Uno, Y., Okada, T., Ikeda, M., Mori, D., Inada, T., Iwata, N., Ozaki, N., 2016. Identification of a rare variant in CHD8 that contributes to schizophrenia and autism spectrum disorder susceptibility. Schizophr. Res. 178, 104–106. https://doi.org/10.1016/j.schres.2016.08.023

Klingler, E., Martin, P.-M., Garcia, M., Moreau-Fauvarque, C., Falk, J., Chareyre, F., Giovannini, M., Chédotal, A., Girault, J.-A., Goutebroze, L., 2015. The cytoskeleton-associated protein SCHIP1 is involved in axon guidance, and is required for piriform cortex and anterior commissure development. Development 142, 2026–2036. https://doi.org/10.1242/dev.119248

Ko, J., 2012. The leucine-rich repeat superfamily of synaptic adhesion molecules: LRRTMs and Slitrks. Mol. Cells 34, 335–340. https://doi.org/10.1007/s10059-012-0113-3

Krumm, N., O’Roak, B.J., Shendure, J., Eichler, E.E., 2014. A de novo convergence of autism genetics and molecular neuroscience. Trends Neurosci. 37, 95–105. https://doi.org/10.1016/j.tins.2013.11.005

Kvarnung, M., Shahsavani, M., Taylan, F., Moslem, M., Breeuwsma, N., Laan, L., Schuster, J., Jin, Z., Nilsson, D., Lieden, A., Anderlid, B.-M., Nordenskjöld, M., Syk Lundberg, E., Birnir, B., Dahl, N., Nordgren, A., Lindstrand, A., Falk, A., 2019. Ataxia in Patients With Bi-Allelic NFASC Mutations and Absence of Full-Length NF186. Front. Genet. 10. https://doi.org/10.3389/fgene.2019.00896

Lee, S.-K., Jurata, L.W., Nowak, R., Lettieri, K., Kenny, D.A., Pfaff, S.L., Gill, G.N., 2005. The LIM domain-only protein LMO4 is required for neural tube closure. Mol. Cell. Neurosci. 28, 205–214. https://doi.org/10.1016/j.mcn.2004.04.010

Lerner, V., McCaffery, P.J.A., Ritsner, M.S., 2016. Targeting Retinoid Receptors to Treat Schizophrenia: Rationale and Progress to Date. CNS Drugs 30, 269–280. https://doi.org/10.1007/s40263-016-0316-9

Li, H., Zhu, Y., Morozov, Y.M., Chen, X., Page, S.C., Rannals, M.D., Maher, B.J., Rakic, P., 2019. Disruption of TCF4 regulatory networks leads to abnormal cortical development and mental disabilities. Mol. Psychiatry 24, 1235–1246. https://doi.org/10.1038/s41380-019-0353-0

Li, S., DeLisi, L.E., McDonough, S.I., 2021. Rare germline variants in individuals diagnosed with schizophrenia within multiplex families. Psychiatry Res. 303, 114038. https://doi.org/10.1016/j.psychres.2021.114038

Li, Z., Chen, J., Yu, H., He, L., Xu, Y., Zhang, D., Yi, Q., Li, C., Li, X., Shen, J., Song, Z., Ji, W., Wang, M., Zhou, J., Chen, B., Liu, Y., Wang, J., Wang, P., Yang, P., Wang, Q., Feng, G., Liu, B., Sun, W., Li, B., He, G., Li, Weidong, Wan, C., Xu, Q., Li, Wenjin, Wen, Z., Liu, K., Huang, F., Ji, J., Ripke, S., Yue, W., Sullivan, P.F., O’Donovan, M.C., Shi, Y., 2017. Genome-wide association analysis identifies 30 new susceptibility loci for schizophrenia. Nat. Genet. 49, 1576–1583. https://doi.org/10.1038/ng.3973

Logan, C. V, Szabadkai, G., Sharpe, J.A., Parry, D.A., Torelli, S., Childs, A.-M., Kriek, M., Phadke, R., Johnson, C.A., Roberts, N.Y., Bonthron, D.T., Pysden, K.A., Whyte, T., Munteanu, I., Foley, A.R., Wheway, G., Szymanska, K., Natarajan, S., Abdelhamed, Z.A., Morgan, J.E., Roper, H., Santen, G.W.E., Niks, E.H., van der Pol, W.L., Lindhout, D., Raffaello, A., De Stefani, D., den Dunnen, J.T., Sun, Y., Ginjaar, I., Sewry, C.A., Hurles, M., Rizzuto, R., Duchen, M.R., Muntoni, F., Sheridan, E., 2014. Loss-of-function mutations in MICU1 cause a brain and muscle disorder linked to primary alterations in mitochondrial calcium signaling. Nat. Genet. 46, 188–193. https://doi.org/10.1038/ng.2851

Ludwig, K.U., Mattheisen, M., Mühleisen, T.W., Roeske, D., Schmäl, C., Breuer, R., Schulte-Körne, G., Müller-Myhsok, B., Nöthen, M.M., Hoffmann, P., Rietschel, M., Cichon, S., 2009. Supporting evidence for LRRTM1 imprinting effects in schizophrenia. Mol. Psychiatry 14, 743–745. https://doi.org/10.1038/mp.2009.28

Ma, C., Gu, C., Huo, Y., Li, X., Luo, X.-J., 2018. The integrated landscape of causal genes and pathways in schizophrenia. Transl. Psychiatry 8, 67. https://doi.org/10.1038/s41398-018-0114-x

Ma, L., Semick, S.A., Chen, Q., Li, C., Tao, R., Price, A.J., Shin, J.H., Jia, Y., Brandon, N.J., Cross, A.J., Hyde, T.M., Kleinman, J.E., Jaffe, A.E., Weinberger, D.R., Straub, R.E., Consortium, T.B., 2020. Schizophrenia risk variants influence multiple classes of transcripts of sorting nexin 19 (SNX19). Mol. Psychiatry 25, 831–843. https://doi.org/10.1038/s41380-018-0293-0

Maeta, K., Edamatsu, H., Nishihara, K., Ikutomo, J., Bilasy, S.E., Kataoka, T., 2016. Crucial Role of Rapgef2 and Rapgef6, a Family of Guanine Nucleotide Exchange Factors for Rap1 Small GTPase, in Formation of Apical Surface Adherens Junctions and Neural Progenitor Development in the Mouse Cerebral Cortex. eneuro 3, ENEURO.0142-16.2016. https://doi.org/10.1523/ENEURO.0142-16.2016

McCarthy, S.E., Gillis, J., Kramer, M., Lihm, J., Yoon, S., Berstein, Y., Mistry, M., Pavlidis, P., Solomon, R., Ghiban, E., Antoniou, E., Kelleher, E., O’Brien, C., Donohoe, G., Gill, M., Morris, D.W., McCombie, W.R., Corvin, A., 2014. De novo mutations in schizophrenia implicate chromatin remodeling and support a genetic overlap with autism and intellectual disability. Mol. Psychiatry 19, 652–658. https://doi.org/10.1038/mp.2014.29

Meffre, D., Grenier, J., Bernard, S., Courtin, F., Dudev, T., Shackleford, G., Jafarian-Tehrani, M., Massaad, C., 2014. Wnt and lithium: a common destiny in the therapy of nervous system pathologies? Cell. Mol. Life Sci. 71, 1123–1148. https://doi.org/10.1007/s00018-013-1378-1

Mi, S., Miller, R.H., Lee, X., Scott, M.L., Shulag-Morskaya, S., Shao, Z., Chang, J., Thill, G., Levesque, M., Zhang, M., Hession, C., Sah, D., Trapp, B., He, Z., Jung, V., McCoy, J.M., Pepinsky, R.B., 2005. LINGO-1 negatively regulates myelination by oligodendrocytes. Nat. Neurosci. 8, 745–751. https://doi.org/10.1038/nn1460

Michaelovsky, E., Carmel, M., Frisch, A., Salmon-Divon, M., Pasmanik-Chor, M., Weizman, A., Gothelf, D., 2019. Risk gene-set and pathways in 22q11.2 deletion-related schizophrenia: a genealogical molecular approach. Transl. Psychiatry 9, 15. https://doi.org/10.1038/s41398-018-0354-9

Murphy, C.E., Walker, A.K., Weickert, C.S., 2021. Neuroinflammation in schizophrenia: the role of nuclear factor kappa B. Transl. Psychiatry 11, 528. https://doi.org/10.1038/s41398-021-01607-0

Narayan, S., Head, S.R., Gilmartin, T.J., Dean, B., Thomas, E.A., 2009. Evidence for disruption of sphingolipid metabolism in schizophrenia. J. Neurosci. Res. 87, 278–288. https://doi.org/10.1002/jnr.21822

Nielsen, S.M., Toftdahl, N.G., Nordentoft, M., Hjorthøj, C., 2017. Association between alcohol, cannabis, and other illicit substance abuse and risk of developing schizophrenia: a nationwide population based register study. Psychol. Med. 47, 1668–1677. https://doi.org/10.1017/S0033291717000162

O’Dushlaine, C., Kenny, E., Heron, E., Donohoe, G., Gill, M., Morris, D., Corvin, A., Consortium, T.I.S., 2011. Molecular pathways involved in neuronal cell adhesion and membrane scaffolding contribute to schizophrenia and bipolar disorder susceptibility. Mol. Psychiatry 16, 286–292. https://doi.org/10.1038/mp.2010.7

Ono, K., Tomasiewicz, H., Magnuson, T., Rutishauser, U., 1994. N-CAM mutation inhibits tangential neuronal migration and is phenocopied by enzymatic removal of polysialic acid. Neuron 13, 595–609. https://doi.org/10.1016/0896-6273(94)90028-0

Perocchi, F., Gohil, V.M., Girgis, H.S., Bao, X.R., McCombs, J.E., Palmer, A.E., Mootha, V.K., 2010. MICU1 encodes a mitochondrial EF hand protein required for Ca2+ uptake. Nature 467, 291–296. https://doi.org/10.1038/nature09358

Phan, B.N., Bohlen, J.F., Davis, B.A., Ye, Z., Chen, H.-Y., Mayfield, B., Sripathy, S.R., Cerceo Page, S., Campbell, M.N., Smith, H.L., Gallop, D., Kim, H., Thaxton, C.L., Simon, J.M., Burke, E.E., Shin, J.H., Kennedy, A.J., Sweatt, J.D., Philpot, B.D., Jaffe, A.E., Maher, B.J., 2020. A myelin-related transcriptomic profile is shared by Pitt–Hopkins syndrome models and human autism spectrum disorder. Nat. Neurosci. 23, 375–385. https://doi.org/10.1038/s41593-019-0578-x

Qin, Z., Zhang, L., Cruz, S.A., Stewart, A.F.R., Chen, H.-H., 2020. Activation of tyrosine phosphatase PTP1B in pyramidal neurons impairs endocannabinoid signaling by tyrosine receptor kinase trkB and causes schizophrenia-like behaviors in mice. Neuropsychopharmacology 45, 1884–1895. https://doi.org/10.1038/s41386-020-0755-3

Qin, Z., Zhou, X., Pandey, N.R., Vecchiarelli, H.A., Stewart, C.A., Zhang, X., Lagace, D.C., Brunel, J.M., Béïque, J.-C., Stewart, A.F.R., Hill, M.N., Chen, H.-H., 2015. Chronic Stress Induces Anxiety via an Amygdalar Intracellular Cascade that Impairs Endocannabinoid Signaling. Neuron 85, 1319–1331. https://doi.org/10.1016/j.neuron.2015.02.015

Quednow, B.B., Brzózka, M.M., Rossner, M.J., 2014. Transcription factor 4 (TCF4) and schizophrenia: integrating the animal and the human perspective. Cell. Mol. Life Sci. 71, 2815–2835. https://doi.org/10.1007/s00018-013-1553-4

Raghu, P., Joseph, A., Krishnan, H., Singh, P., Saha, S., 2019. Phosphoinositides: Regulators of Nervous System Function in Health and Disease. Front. Mol. Neurosci. 12. https://doi.org/10.3389/fnmol.2019.00208

Reay, W.R., Atkins, J.R., Quidé, Y., Carr, V.J., Green, M.J., Cairns, M.J., 2020. Polygenic disruption of retinoid signalling in schizophrenia and a severe cognitive deficit subtype. Mol. Psychiatry 25, 719–731. https://doi.org/10.1038/s41380-018-0305-0

Roussos, P., 2012. Molecular and Genetic Evidence for Abnormalities in the Nodes of Ranvier in Schizophrenia. Arch. Gen. Psychiatry 69, 7. https://doi.org/10.1001/archgenpsychiatry.2011.110

Shi, R.-Y., Zhu, S.-H., Li, V., Gibson, S.B., Xu, X.-S., Kong, J.-M., 2014. BNIP3 Interacting with LC3 Triggers Excessive Mitophagy in Delayed Neuronal Death in Stroke. CNS Neurosci. Ther. 20, 1045–1055. https://doi.org/10.1111/cns.12325

Shimoda, Y., Watanabe, K., 2009. Contactins. Cell Adh. Migr. 3, 64–70. https://doi.org/10.4161/cam.3.1.7764

Silva, J.-P., Lelianova, V.G., Ermolyuk, Y.S., Vysokov, N., Hitchen, P.G., Berninghausen, O., Rahman, M.A., Zangrandi, A., Fidalgo, S., Tonevitsky, A.G., Dell, A., Volynski, K.E., Ushkaryov, Y.A., 2011. Latrophilin 1 and its endogenous ligand Lasso/teneurin-2 form a high-affinity transsynaptic receptor pair with signaling capabilities. Proc. Natl. Acad. Sci. 108, 12113–12118. https://doi.org/10.1073/pnas.1019434108

Srivastava, S., Engels, H., Schanze, I., Cremer, K., Wieland, T., Menzel, M., Schubach, M., Biskup, S., Kreiß, M., Endele, S., Strom, T.M., Wieczorek, D., Zenker, M., Gupta, S., Cohen, J., Zink, A.M., Naidu, S., 2016. Loss-of-function variants in HIVEP2 are a cause of intellectual disability. Eur. J. Hum. Genet. 24, 556–561. https://doi.org/10.1038/ejhg.2015.151

Stanta, J.L., Saldova, R., Struwe, W.B., Byrne, J.C., Leweke, F.M., Rothermund, M., Rahmoune, H., Levin, Y., Guest, P.C., Bahn, S., Rudd, P.M., 2010. Identification of N-Glycosylation Changes in the CSF and Serum in Patients with Schizophrenia. J. Proteome Res. 9, 4476–4489. https://doi.org/10.1021/pr1002356

Steinfeld, H., Cho, M.T., Retterer, K., Person, R., Schaefer, G.B., Danylchuk, N., Malik, S., Wechsler, S.B., Wheeler, P.G., van Gassen, K.L.I., Terhal, P.A., Verhoeven, V.J.M., van Slegtenhorst, M.A., Monaghan, K.G., Henderson, L.B., Chung, W.K., 2016. Mutations in HIVEP2 are associated with developmental delay, intellectual disability, and dysmorphic features. Neurogenetics 17, 159–164. https://doi.org/10.1007/s10048-016-0479-z

Sugathan, A., Biagioli, M., Golzio, C., Erdin, S., Blumenthal, I., Manavalan, P., Ragavendran, A., Brand, H., Lucente, D., Miles, J., Sheridan, S.D., Stortchevoi, A., Kellis, M., Haggarty, S.J., Katsanis, N., Gusella, J.F., Talkowski, M.E., 2014. CHD8 regulates neurodevelopmental pathways associated with autism spectrum disorder in neural progenitors. Proc. Natl. Acad. Sci. 111, E4468–E4477. https://doi.org/10.1073/pnas.1405266111

Takashima, N., Odaka, Y.S., Sakoori, K., Akagi, T., Hashikawa, T., Morimura, N., Yamada, K., Aruga, J., 2011. Impaired Cognitive Function and Altered Hippocampal Synapse Morphology in Mice Lacking Lrrtm1, a Gene Associated with Schizophrenia. PLoS One 6, e22716. https://doi.org/10.1371/journal.pone.0022716

Talkowski, M.E., Rosenfeld, J.A., Blumenthal, I., Pillalamarri, V., Chiang, C., Heilbut, A., Ernst, C., Hanscom, C., Rossin, E., Lindgren, A.M., Pereira, S., Ruderfer, D., Kirby, A., Ripke, S., Harris, D.J., Lee, J.-H., Ha, K., Kim, H.-G., Solomon, B.D., Gropman, A.L., Lucente, D., Sims, K., Ohsumi, T.K., Borowsky, M.L., Loranger, S., Quade, B., Lage, K., Miles, J., Wu, B.-L., Shen, Y., Neale, B., Shaffer, L.G., Daly, M.J., Morton, C.C., Gusella, J.F., 2012. Sequencing Chromosomal Abnormalities Reveals Neurodevelopmental Loci that Confer Risk across Diagnostic Boundaries. Cell 149, 525–537. https://doi.org/10.1016/j.cell.2012.03.028

Tanaka, S., Terada, K., Nohno, T., 2011. Canonical Wnt signaling is involved in switching from cell proliferation to myogenic differentiation of mouse myoblast cells. J. Mol. Signal. 6, 12. https://doi.org/10.1186/1750-2187-6-12

Teixeira, J.R., Szeto, R.A., Carvalho, V.M.A., Muotri, A.R., Papes, F., 2021. Transcription factor 4 and its association with psychiatric disorders. Transl. Psychiatry 11, 19. https://doi.org/10.1038/s41398-020-01138-0

Um, J.W., Choi, T.-Y., Kang, H., Cho, Y.S., Choii, G., Uvarov, P., Park, D., Jeong, D., Jeon, S., Lee, D., Kim, H., Lee, S.-H., Bae, Y.-C., Choi, S.-Y., Airaksinen, M.S., Ko, J., 2016. LRRTM3 Regulates Excitatory Synapse Development through Alternative Splicing and Neurexin Binding. Cell Rep. 14, 808–822. https://doi.org/10.1016/j.celrep.2015.12.081

Valvezan, A.J., Klein, P.S., 2012. GSK-3 and Wnt Signaling in Neurogenesis and Bipolar Disorder. Front. Mol. Neurosci. 5. https://doi.org/10.3389/fnmol.2012.00001

Veeraval, L., O’Leary, C.J., Cooper, H.M., 2020. Adherens Junctions: Guardians of Cortical Development. Front. Cell Dev. Biol. 8. https://doi.org/10.3389/fcell.2020.00006

Vieira, N., Rito, T., Correia-Neves, M., Sousa, N., 2021. Sorting Out Sorting Nexins Functions in the Nervous System in Health and Disease. Mol. Neurobiol. 58, 4070–4106. https://doi.org/10.1007/s12035-021-02388-9

Wang, P., Mokhtari, R., Pedrosa, E., Kirschenbaum, M., Bayrak, C., Zheng, D., Lachman, H.M., 2017. CRISPR/Cas9-mediated heterozygous knockout of the autism gene CHD8 and characterization of its transcriptional networks in cerebral organoids derived from iPS cells. Mol. Autism 8, 11. https://doi.org/10.1186/s13229-017-0124-1

Wedel, M., Fröb, F., Elsesser, O., Wittmann, M.-T., Lie, D.C., Reis, A., Wegner, M., 2020. Transcription factor Tcf4 is the preferred heterodimerization partner for Olig2 in oligodendrocytes and required for differentiation. Nucleic Acids Res. 48, 4839–4857. https://doi.org/10.1093/nar/gkaa218

Wu, L.-C., 2002. ZAS: C2H2 Zinc Finger Proteins Involved in Growth and Development. Gene Expr. 10, 137–152. https://doi.org/10.3727/000000002783992479

Xu, B., Roos, J.L., Levy, S., van Rensburg, E.J., Gogos, J.A., Karayiorgou, M., 2008. Strong association of de novo copy number mutations with sporadic schizophrenia. Nat. Genet. 40, 880–885. https://doi.org/10.1038/ng.162

Xu, B., Woodroffe, A., Rodriguez-Murillo, L., Roos, J.L., van Rensburg, E.J., Abecasis, G.R., Gogos, J.A., Karayiorgou, M., 2009. Elucidating the genetic architecture of familial schizophrenia using rare copy number variant and linkage scans. Proc. Natl. Acad. Sci. 106, 16746–16751. https://doi.org/10.1073/pnas.0908584106

Xu, C., Aragam, N., Li, X., Villla, E.C., Wang, L., Briones, D., Petty, L., Posada, Y., Arana, T.B., Cruz, G., Mao, C., Camarillo, C., Su, B. Bin, Escamilla, M.A., Wang, K., 2013. BCL9 and C9orf5 Are Associated with Negative Symptoms in Schizophrenia: Meta-Analysis of Two Genome-Wide Association Studies. PLoS One 8, e51674. https://doi.org/10.1371/journal.pone.0051674

Zhang, Z., Xu, X., Zhang, Y., Zhou, J., Yu, Z., He, C., 2009. LINGO-1 Interacts with WNK1 to Regulate Nogo-induced Inhibition of Neurite Extension. J. Biol. Chem. 284, 15717–15728. https://doi.org/10.1074/jbc.M808751200

Zhao, Z., Webb, B.T., Jia, P., Bigdeli, T.B., Maher, B.S., van den Oord, E., Bergen, S.E., Amdur, R.L., O’Neill, F.A., Walsh, D., Thiselton, D.L., Chen, X., Pato, C.N., Riley, B.P., Kendler, K.S., Fanous, A.H., 2013. Association Study of 167 Candidate Genes for Schizophrenia Selected by a Multi-Domain Evidence-Based Prioritization Algorithm and Neurodevelopmental Hypothesis. PLoS One 8, e67776. https://doi.org/10.1371/journal.pone.0067776

Zhou, J., Ma, C., Wang, K., Li, X., Jian, X., Zhang, H., Yuan, J., Yin, J., Chen, J., Shi, Y., 2020. Identification of rare and common variants in BNIP3L: a schizophrenia susceptibility gene. Hum. Genomics 14, 16. https://doi.org/10.1186/s40246-020-00266-4
